# The 40 Hz auditory steady state response is associated with antipsychotic treatment outcome in acute patients with schizophrenia

**DOI:** 10.64898/2026.01.26.26344882

**Authors:** Marco de Pieri, Vincent Rochas, Cyril Petignat, Geraldina Molo, Nadege Bertrand, Dafni Apostolopoulou, Michel Godel, Matthias Kirschner, Stefan Kaiser

## Abstract

**Background:** Prediction of response to antipsychotic medications remains elusive, and a biomarker assisting in treatment selection would drastically improve prognosis. The 40 Hz auditory steady state response (ASSR) is an EEG biomarker, mirroring the GABA-glutamate signaling and the excitation/inhibition balance, consistently been reported to be impaired in schizophrenia, on, with inconsistent evidence of an association with specific symptoms.

**Methods:** N=69 schizophrenia inpatients with an acute psychotic episode underwent an EEG recording to assess event related spectral perturbation (ERSP), intertrial phase coherence (ITC) and phase amplitude coupling (PAC) during the ASSR task, aimed to assess their relationship with response to antipsychotics and with positive, negative, disorganized, excited and depressive symptoms. Moreover, patients were compared with controls (N=36), to delineate schizophrenia acute phase ASSR dynamics.

**Results:** Responders to treatment showed a decreased 40 Hz ERSP in both the early (0-0.2s post-stimulus; P=0.0013; d=-0.936) and late (0-2-1.2s post-stimulus; P=0.0022; d=-0.932) time windows compared to non-responders. Using logistic regression and bootstrap optimism correction, ERSP classified the two groups with 70% accuracy. Responders but not non-responders showed a reduced ERSP compared to controls (P=0.0211; d=-0.558). Patients had reduced early ITPC (P=0.0001; d=-1.015) vs controls. responders compared to non-responders had increased PAC in the early (P=0.0215; d00.65) and in patients vs controls, in both the early (P=0.0002; d=0.57) and the late (P=0.0006; d=0.74) windows. No association emerged between ASSR metrics and symptoms severity.

**Conclusions:** ASSR is a candidate biomarker for antipsychotic treatment personalization. Only responders to treatment presented a significant gamma-band impairment, in line with previous literature on stabilized outpatients, but not non-responders, indicating that a distinct neurobiology could exist.

## INTRODUCTION

Antipsychotic medications (AP) represent the mainstay in the treatment of SCZ, but response rate ranges from 47% for individuals who have received prior treatment to 66% for AP-naive individuals^1^, while 1-year discontinuation rates from 32% to 74%, due to poor tolerability and/or lack of efficacy^2^. Interindividual variability in response to AP is large and unpredictable, giving rise to the practice of prescription by trial and error, often requiring multiple attempts with different molecules, dose adjustments and drug combinations. Predictors of AP response remains elusive: neuroimaging (both structural and functional), genetics and molecular biology did not produce a biomarker for AP selection in clinical practice^3, 4^, which would be of the utmost relevance to deliver timely and effective treatments^5, 6^.

The glutamatergic theory of schizophrenia posits the hypoactivity of the N-methyl-D-aspartate (NMDA) glutamate receptors located on the gamma amino-butyric acid (GABA) parvalbumin-containing interneurons (PV-interneurons) as a core pathogenetic process, leading to an excitation/inhibition (E/I) imbalance of the pyramidal neuron^7, 8^, which in turn also disrupts the dopaminergic mesolimbic and mesocortical neurotransmission^9^.

EEG gamma frequencies are brain oscillations in the 30–100 hz range, originating from the interplay between pyramidal neurons and PV-interneurons, thus linked to the glutamatergic theory and the E/I imbalance. In fact, gamma activity is generated through feedback inhibition on pyramidal neurons by PV-interneurons occurring in all regions^10, 11^; the decay rate of this inhibition, approximately 25 ms, particularly contributes to the generation of 40 Hz oscillations^12^.

Classically both resting state and task related gamma band activity were deemed important for schizophrenia;, however, the literature focusing on the resting state produced mixed, contradictory evidence^13^, while evidence on evoked activity was solid, in particular concerning the auditory steady state response (ASSR). Steady-state responses (SSRs) are evoked oscillatory responses that are entrained to the frequency and phase of temporally modulated stimuli. SSRs reflect the propensity of neurons to oscillate at a particular “resonant” frequency band induced by external periodic stimulation, that in the auditory modality (ASSR) show a peak at 40 Hz^14^.

The classical analysis of the ASSR comprises power (event related spectral perturbation, ERSP), and phase (intertrial phase coherence, ITC) measures, in two time windows: an early phase reflecting event-related potential processes, and late latency sustained responses reflecting rhythmic activity^15^. Kwon et al first investigated 40-Hz ASSRs in patients with schizophrenia, demonstrating both a reduced power and a phase delay at the 40-Hz stimulation ^16^. After 26 years of research on the topic, approximately 40 studies have investigated 40 Hz ASSR in patients with schizophrenia using M/EEG, and the large majority found a reduction of both ITC and ERSP with medium level effect size^14, 17^. Few studies also investigated the association of ASSR parameters with positive and negative symptoms of schizophrenia, with contradictory results.

Another parameter investigated in a minority of study is the phase amplitude coupling (PAC), a neural mechanism where the phase of a slow brainwave rhythmically controls the amplitude of a faster brainwave. PAC accounts for the synchronization of brain functions subtended by theta and gamma frequencies, and it was so far studied in few studies on schizophrenia, often with non-significant results^18-20^, with only one study showing a complex pattern of increase and decrease in different brain regions, in patients vs controls^21^.

The primary objective of our study was to assess the role of the 40 Hz auditory steady state response as a predictor of the response to antipsychotic medications.

We expected that response to antipsychotic medications was related to lower ASSR power (ERSP) and phase synchronization (ITC), in line with the glutamatergic dysfunction and the E/I imbalance ASSR can mirror. Hypothesizing that impaired gamma band activity is a direct expression of the core pathophysiology process of schizophrenia, we expected that it also represented the substrate for AP action: thus, the larger the impairment, the more likely that APs can exert a significant effect.

The secondary objective of the study was to explore if ASSR parameters, besides being related to schizophrenia as a unitary entity, also correlated with positive, negative, disorganized, excited, and depressive symptoms dimensions, an outcome so far less explored and with unconclusive findings. The tertiary objective of the study was to assess ASSR dynamics in acute patients with schizophrenia, by comparing them with healthy controls. We expected to find an impairment of both ERSP and ITC, similarly to the well replicated finding in stabilized outpatients, however with a larger effect size, given the more severe psychopathology.

Concerning PAC, the investigation was purely explorative, given the unconclusive previous results; we considered that both a lack or an excess of theta-gamma coordination were potentially hindering effective information processing in the auditory regions, leading to aberrant salience, thus influencing treatment response and causing positive and negative symptoms.

## METHODS AND MATERIALS

We recruited 69 adult inpatients with schizophrenia during an acute psychotic episode and 31 matched healthy controls. Patients were evaluated upon admission and after six weeks of antipsychotic treatment; response was defined as a ≥50% reduction in PANSS total score. EEG was recorded during a 40-Hz Auditory Steady-State Response (ASSR) paradigm. Preprocessing involved band-pass filtering (1-80 Hz), artifact removal via ICA, and interpolation. Time-frequency representations were computed using multitapers, with 40-Hz Event-Related Spectral Perturbation (ERSP) and Inter-Trial Phase Coherence (ITPC) extracted for early (0-300ms) and late (300-1200ms) windows relative to a baseline optimized for maximal contrast. We also computed theta-gamma Phase-Amplitude Coupling (PAC) using the Modulation Index. Group differences (Patients vs. Controls; Responders vs. Non-Responders) were assessed using non-parametric cluster-based permutation tests (10,000 permutations) to control for multiple comparisons. Clinical confounding factors were evaluated using Spearman correlations and Kruskal-Wallis tests (FDR-corrected). To evaluate translational utility, we constructed a binary logistic regression model predicting treatment response based on the mean 40-Hz ERSP from the significant cluster identified in the permutation test. Model performance (AUC, sensitivity, specificity) was internally validated using Harrell’s bootstrap optimism correction (1,000 iterations) to provide bias-corrected estimates of discriminative power. Full details on recruitment, preprocessing, and modeling are provided in **Supplementary methods**.

## RESULTS

### Participants and treatment characteristics

Table 1. shows that socio-demographic and clinical features in controls, responders and non-responders to treatment presented no significant differences, including age, sex, handedness, education, education of the parents, duration of illness, duration of untreated psychosis.

**Table 1:** sociodemographic and clinical features of patients and controls.

|  | **Responders (R)**<br>**N=38** | **Non-Resp (NR)**<br>**N=31** | **Controls (C)**<br>**N=36** | **R vs NR**<br>**(p)** | **R vs NR**<br>**(ES)** | **R vs C**<br>**(p)** | **R vs C**<br>**(ES)** | **NR vs C**<br>**(p)** | **NR vs C**<br>**(ES)** |
| --- | --- | --- | --- | --- | --- | --- | --- | --- | --- |
| **Sex (M)** | 78.9% | 83.9% | 69.4% | Ns | V=0.06 | Ns | V=0.11 | Ns | V=0.17 |
| **Age** | 34.87 ± 10.67 | 32.00 ± 9.96 | 31.56 ± 7.65 | Ns | d=0.28 | Ns | d=0.36 | Ns | d=0.05 |
| **Handedness (right)** | 80.0% | 96.3% | 94.1% | 0.085 | V=0.29 | Ns | V=0.22 | Ns | V=0.22 |
| **Education (years)** | 14.41 ± 3.64 | 14.90 ± 3.84 | 19.25 ± 2.69 | 0.804 | d=-0.13 | **0.00001** | d=-1.51 | **0.00001** | d=-1.33 |
| **Education mother** | 12.81 ± 6.27 | 13.67 ± 7.05 | 16.54 ± 5.45 | 0.621 | d=-0.13 | Ns | d=-0.64 | Ns | d=-0.47 |
| **Education father** | 13.54 ± 6.29 | 14.87 ± 7.11 | 17.15 ± 5.32 | 0.284 | d=-0.20 | Ns | d=-0.63 | Ns | d=-0.37 |
| **Age of onset** | 24.78 ± 8.83 | 22.35 ± 7.32 |  | Ns | d=0.30 |  |  |  |  |
| **Duration of Illness (months)** | 125.59 ± 128.75 | 119.52 ± 111.42 |  | Ns | d=0.05 |  |  |  |  |
| **Duration of untreated psychosis** | 33.03 ± 54.00 | 16.35 ± 29.04 |  | Ns | d=0.37 |  |  |  |  |
| **Number of hospitalizations** | 5.76 ± 5.15 | 6.63 ± 5.34 |  | Ns | d=-0.17 |  |  |  |  |
| **PANSS Total** | 100.53 ± 26.09 | 100.14 ± 25.43 |  | Ns | d=0.02 |  |  |  |  |
| **PANSS Positive** | 15.79 ± 5.11 | 15.37 ± 3.52 |  | Ns | d=0.09 |  |  |  |  |
| **PANSS Negative** | 20.37 ± 7.64 | 21.90 ± 6.50 |  | Ns | d=-0.21 |  |  |  |  |
| **PANSS Disorganized** | 11.61 ± 3.53 | 11.20 ± 2.78 |  | Ns | d=0.13 |  |  |  |  |
| **PANSS Excited** | 15.03 ± 6.28 | 14.20 ± 9.50 |  | Ns | d=0.11 |  |  |  |  |
| **PANSS Depressed** | 6.42 ± 3.14 | 7.20 ± 3.46 |  | Ns | d=-0.24 |  |  |  |  |

When comparing patients and controls, all the above-mentioned socio-demographic variables were assessed, and no differences emerged as a consequence of the matching process, except for the education level (P=0.001; d=1.151).

At the moment of the EEG recording, all the patients had started an antipsychotic medication: treatment characteristics are reported in **table 2**, and no differences emerged between responders and non-responders.

**Table 2:** antipsychotic medications.

|  | **Responders (R)**<br>N=38 | **Non-Responders (NR)**<br>N=31 | **R vs NR (p)** | **R vs NR (ES)** |
| --- | --- | --- | --- | --- |
| **Risperidone equivalents** | 5.67 ± 3.07 | 5.14 ± 2.34 | Ns | d=0.19 |
| **Main AP Category** |  |  | Ns | V=0.11 |
| “Pines” | 14 (36.8%) | 12 (38.7%) | 1.000 | V=0.02 |
| “Dones” | 18 (47.4%) | 13 (41.9%) | 0.808 | V=0.05 |
| *Partial agonists* | 1 (2.6%) | 2 (6.5%) | 0.584 | V=0.09 |
| *First generation* | 3 (7.9%) | 2 (6.5%) | 1.000 | V=0.03 |
| *Amisulpride* | 2 (5.3%) | 1 (3.2%) | 1.000 | V=0.05 |
| **Main AP breakdown** |  |  |  |  |
| *amisulpride* | 2 (5.3%)<br>450.0±212.1 mg | 1 (3.3%)<br>400 mg | Ns | V=0.05 |
| *aripiprazole* | 1 (2.6%)<br>13.3 mg | 1 (3.3%)<br>25 mg | Ns | V=0.02 |
| *brexpiprazole* | 0 (0.0%) | 1 (3.3%)<br>3 mg | Ns | V=0.14 |
| *clozapine* | 6 (15.8%)<br>391.7±149.7 mg | 6 (20.0%)<br>320.8±64.1mg | Ns | V=0.05 |
| *haloperidol* | 3 (7.9%)<br>16.7±5.8 mg | 2 (6.7%)<br>10.5±0.7 mg | Ns | V=0.02 |
| *olanzapine* | 6 (15.8%)<br>15.0±5.5 mg | 5 (16.7%)<br>13.0±5.7 mg | Ns | V=0.01 |
| *paliperidone* | 7 (18.4%)<br>5.9±3.4 mg | 6 (20.0%)<br>4.7±2.3 mg | Ns | V=0.02 |
| *quetiapine* | 2 (5.3%)<br>550.0±353.6 mg | 2 (6.7%)<br>650.0±212.1 mg | Ns | V=0.03 |
| *risperidone* | 11 (28.9%)<br>3.3±1.3 mg | 6 (19.4%)<br>3.8±1.0mg | Ns | V=0.11 |
| **Monotherapy/combined therapy** | 29/9 (76.3/23.7%) | 21/10 (67/33%) | Ns | V=0.18 |
| **Secondary AP breakdown** |  |  |  |  |
| *amisulpride* | 0 (0.0%) | 2 (6.7%)<br>350.0±353.6 mg | Ns | V=0.20 |
| *aripiprazole* | 2 (5.3%)<br>16.6±4.7 mg | 0 (0.0%) | Ns | V=0.15 |
| *cariprazine* | 0 (0.0%) | 2 (6.7%)<br>6.0±0.0 mg | Ns | V=0.20 |
| *haloperidol* | 2 (5.3%)<br>15.0±0.0 mg | 1 (3.3%)<br>5 mg | Ns | V=0.05 |
| *lurasidone* | 0 (0.0%) | 1 (3.3%)<br>120 mg | Ns | V=0.14 |
| *olanzapine* | 0 (0.0%) | 1 (3.3%)<br>5 mg | Ns | V=0.14 |
| *paliperidone* | 1 (2.6%)<br>3.9 mg | 1 (3.3%)<br>6 mg | Ns | V=0.02 |
| *promazine* | 1 (2.6%)<br>150 mg | 1 (3.3%)<br>100.0 mg | Ns | V=0.02 |
| *risperidone* | 3 (7.9%)<br>3.7±0.6 mg | 1 (3.3%)<br>2 mg | Ns | V=0.10 |

Data expressed as daily dose range (mg). \*Defined daily dose method (Leucht et al, 2016). Long-active injectable or therapeutic drug monitoring after 5-6 weeks

### Assessment of confounders on ERSP, ITPC and PAC

All the socio-demographic, clinical and treatment related variables had no significant relationship with the EEG parameters under study, including ERSP, ITC and PAC in early and late gamma and theta windows (**Supplementary table S2**).

### Reduced gamma and theta ERSP and PAC, but not ITC are associated with response to antipsychotics

When comparing responders and non-responders to treatment, clusters of decreased ERSP in the gamma range were observed, involving both the early (t=-3.831; P=0.0013; d=-0.936) and the late phase (t=-3.816; P=0.0022; d=-0.932). In both phases, decreased ERSP involved a wide cluster of fronto-central electrodes (N_electrodes_=14), expanding quasi-symmetrically on both hemispheres (**fig. 1**). In the theta band, responders to treatment presented a reduced ERSP in both the early (t=-2.773, P=0.0472, d=-0.677) and the late phase (t=-2.766, P=0.029, d=-0.676) compared to non-responders, localized to a cluster of right hemisphere electrodes (N_electrodes_=3) in the early phase, and in a central-right cluster (N_electrodes_=4) in the late phase.

**Figure 1:**
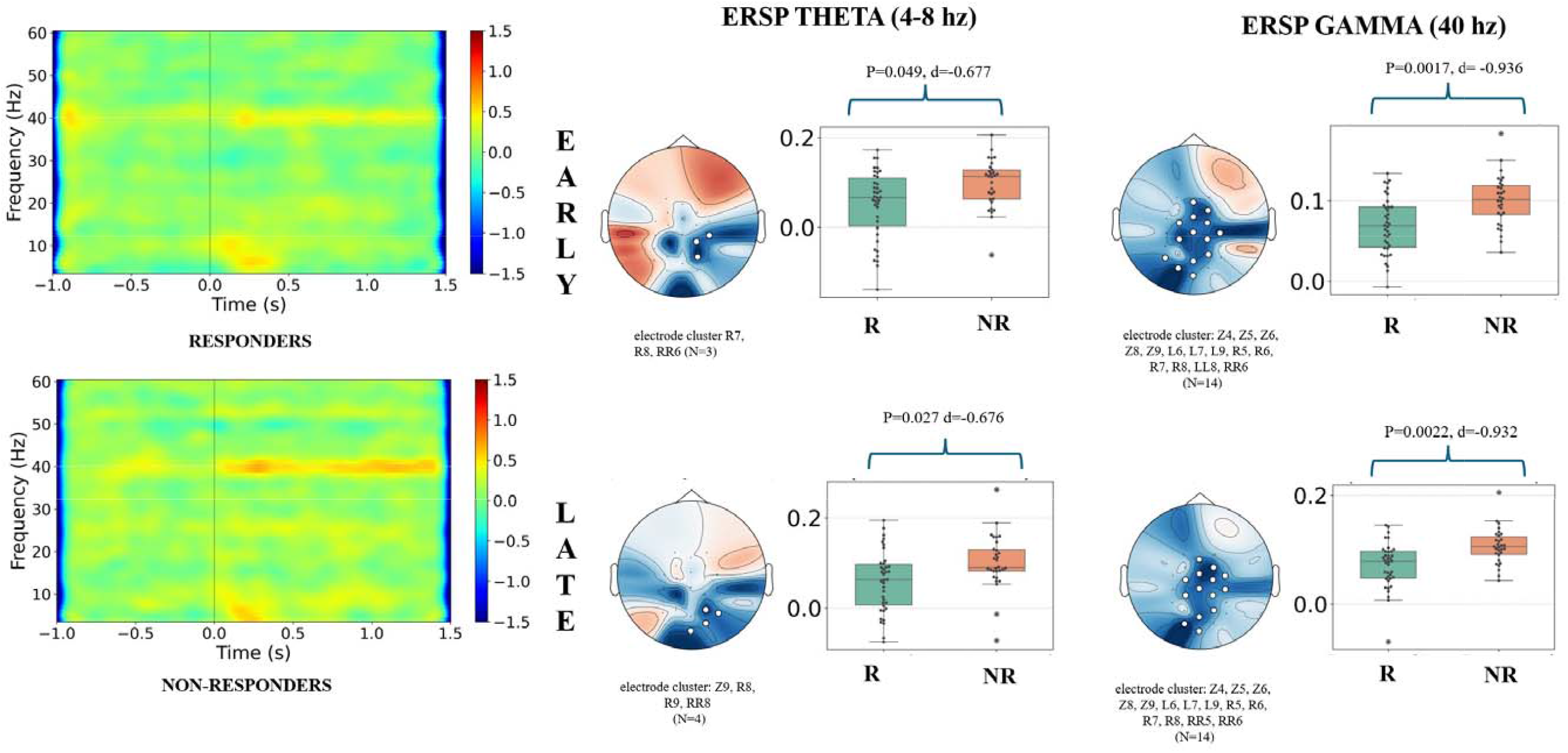
ERSP comparisons between responders and non-responders

Responders vs non-responders to treatment showed an increased PAC in the early window (t= 3.406; P=0.021; d=0.65), in a small cluster of fronto-central left electrodes (N=3), while no differences emerged in the late window (**fig. 4**).

We observed no differences in ITPC in responders vs non responders to treatment, when considering gamma and theta activity in both the early and the late phase.

### ERSP gamma is a candidate biomarker for response to antipsychotics

Using logistic regression, ERSP values were classifying responders and non-responders to treatment with a sensitivity of 79% in the early and 84% in the late window, a specificity of 61% in both the early and the late window and an accuracy of 71% in the early and 74% in the late, yielding to an AUC of 0.73 in the early and 0.76 in the late window.

Applying bootstrap optimism correction only slightly affected results of sensitivity (early ERSP_corrected_: 74%; late ERSP_corrected_: 80%), specificity (early ERSP_corrected_: 57%; late ERSP_corrected_: 57%), accuracy (early_corrected_: 70%; late_corrected_: 67%) and AUC (early ERSP_corrected_: 0.73; late ERSP_corrected_: 0.75).

### Only responders to treatment had reduced theta and gamma ERSP compared to controls

No differences between patients and controls were observed in ERSP gamma, nor between non-responders and controls; however, responders showed a cluster of reduced late ERSP gamma (t=-2.398; P=0.0211; d=-0.558) in a central cluster of electrodes (N_electrodes_=7), asymmetric to the left, compared to controls (**fig. 2**).

**Figure 2:**
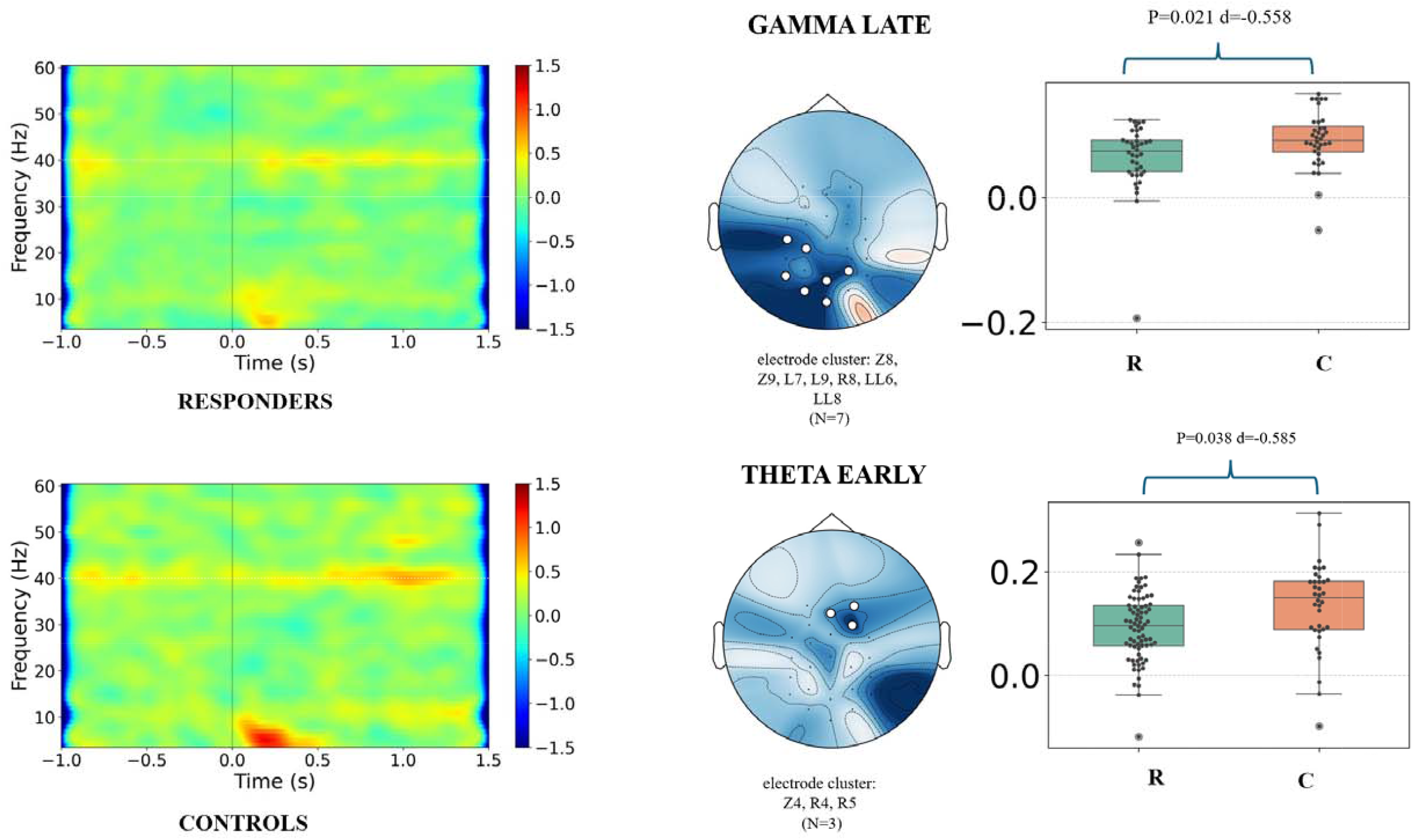
ERSP comparison between patients and controls

A reduced early ERSP theta was also observed when comparing patients with controls (t=-2.887, P=0.039, d=-0.585), in a small frontal-right cluster (N_electrodes_=3); the association was likely driven by responders (t=-2.789, P=0.034, d=-0.649), since no significant results was observed for non-responders vs controls (**fig. 2**). See **supplementary table S3** for full results, also including negative findings.

### Acute patients have reduced theta ITPC and increased PAC compared to controls

When comparing patients vs controls, the early theta ITPC was reduced (t=-5.006; P=0.0001; d=-1.015), in a wide frontal-central cluster of electrode, symmetrically expanding to the left and the right hemispheres (N_electrodes_ =23), a result equally driven by responders (t=-4.634; P=0.0001; d=-1.078) and non-responders (t=-4.312; P=0.0003; d=-1.057) to treatment (**fig. 3**). No differences between patients and control emerged for ITPC theta in the late phase, nor between non-responders and controls, however responders to treatment also showed a reduction in ITPC theta in that window (t=-2.046; P=0.0236; d=-0.476; **fig. 3**) fronto-centrally (N_electrodes_ =9). No significant differences emerged in gamma ITPC between all patients, responders, non-responders and controls. See **supplementary table S4** for full results, also including negative findings.

**Figure 3.**
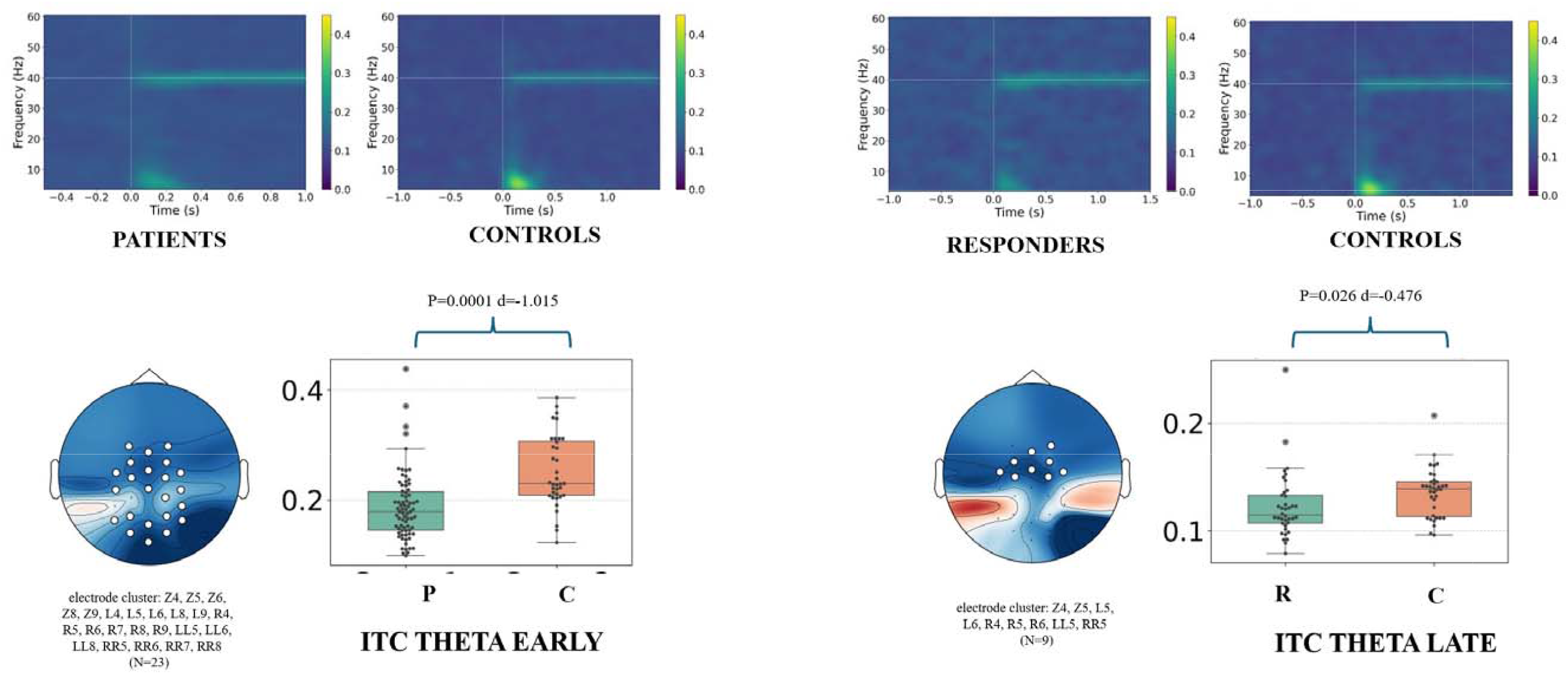
ITPC comparisons between responders, non-responders and controls

**Figure 4:**
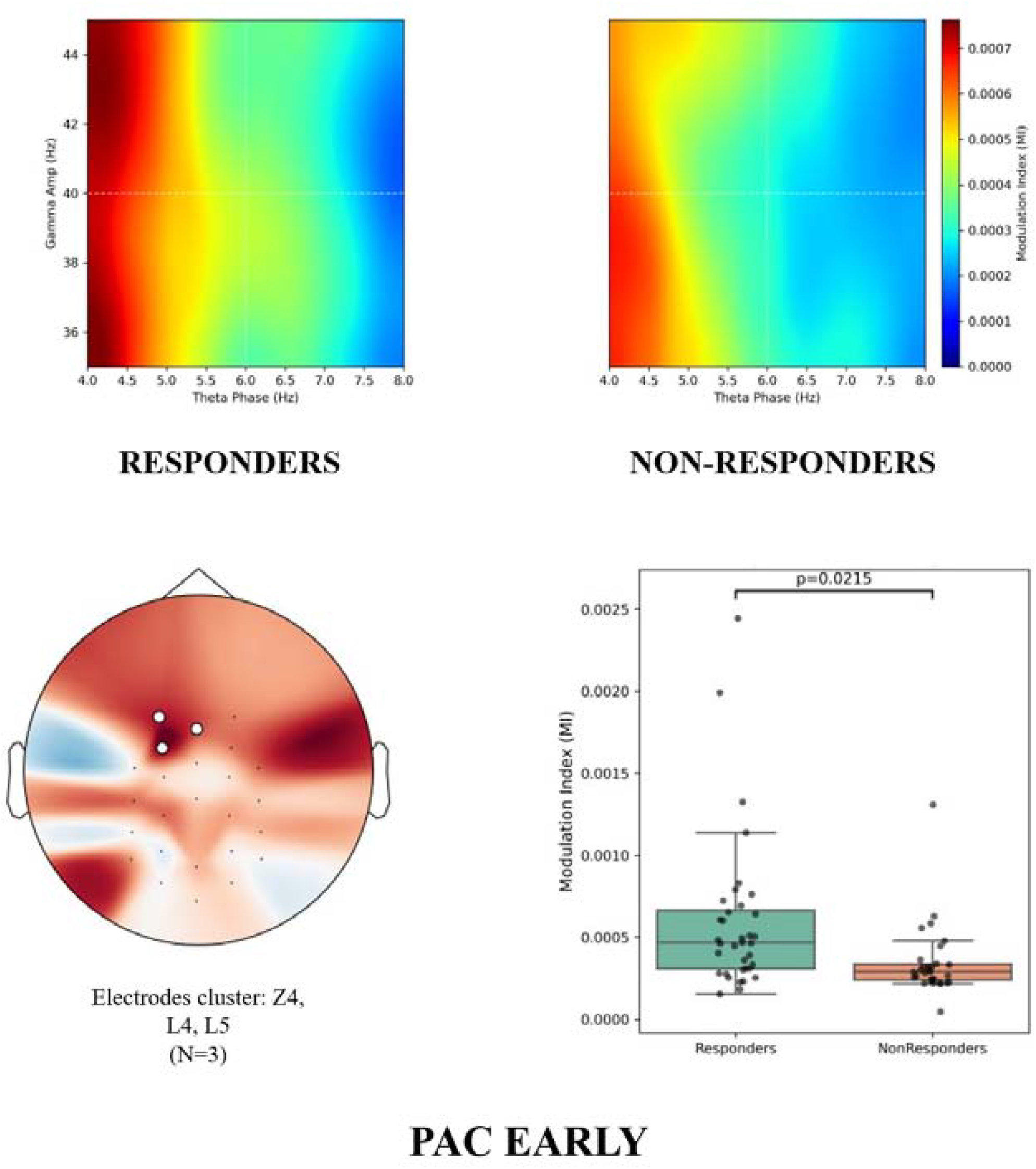
PAC in responders vs non-responders to treatment

Patients compared to controls presented a significant increase in the theta-gamma phase amplitude coupling, in both the early (t= 3.778; P=0.00020; d=0.57) and in the late (t= 4.55; P=0.00060; d=0.74) windows. Early windows difference involved a large cluster (N_electrodes_ =17) of electrodes expanding on both hemispheres, while a similarly distributed cluster emerged from late window (N_electrodes_ =15) (**fig. 5**). We noticed that in the early window only responders differed from controls with a cluster of electrodes with a centro-frontal distribution, slightly asymmetric to the left (t= 3.527; P=0.0003; d=0.822; N_electrodes_=14), while non-responders did not significantly differ from controls. In the late window both responders and non-responders presented a significant increase compared to controls: for the former group differences localized to a right-central cluster (t=2.926; P=0.0078; d=0.68; N_electrodes_=5) and a right-frontal cluster (t=2.933; P=0.0488; d=0.683; N_electrode_s=2) while for the latter group in a central cluster slightly asymmetric to the left (t=3.406; P=0.0135; d=0.883; N_electrodes_=4). See **supplementary table S5** for full results, also including negative findings.

**Figure 5.**
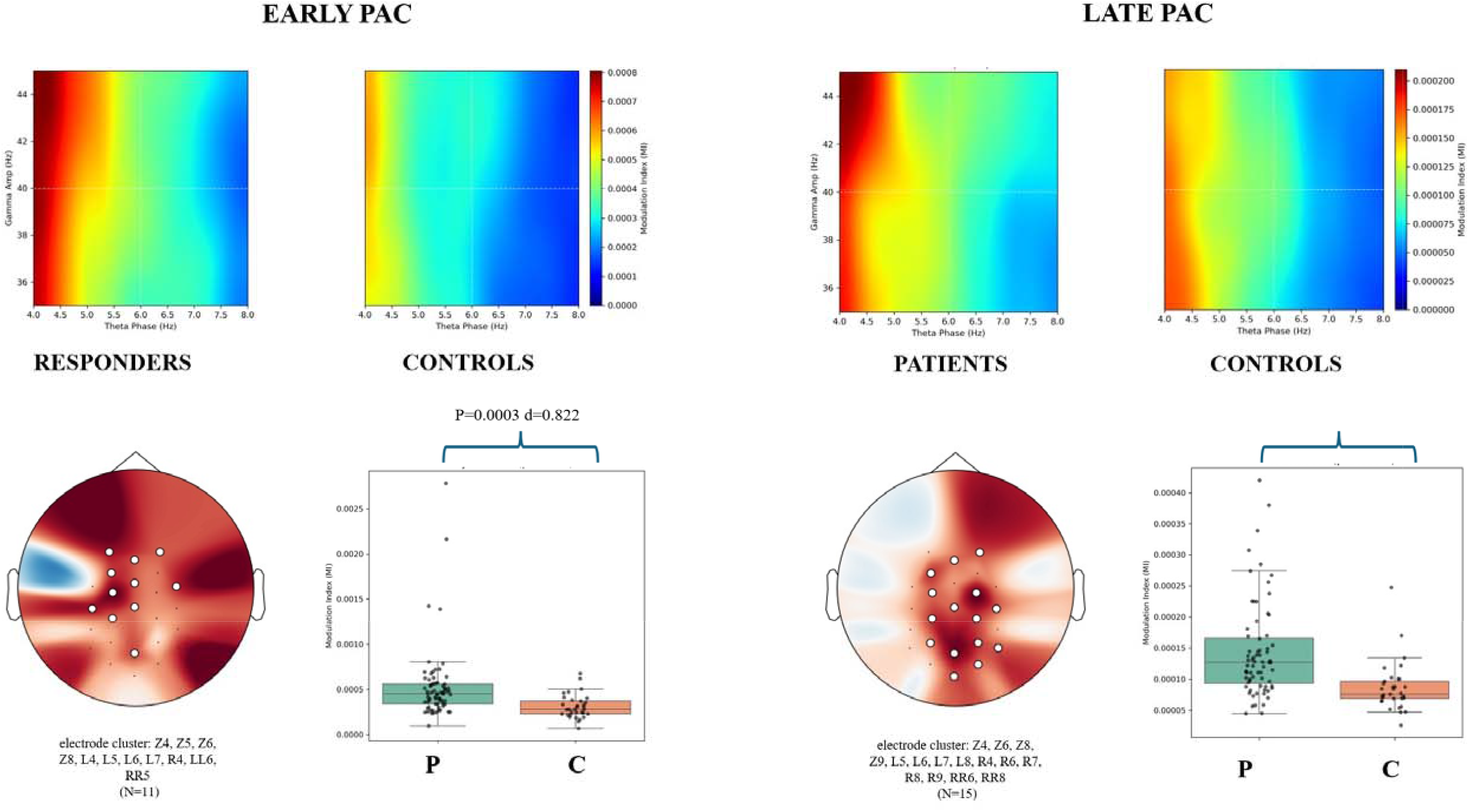
PAC in patients vs controls

### ERSP, ITPC and PAC showed no association with symptoms severity

ERSP and ITC, including theta and gamma band and the early and late windows, did not correlate with the severity of positive, negative, disorganized, excited and depressed symptoms. Similarly, PAC values had no associations with symptoms severity in any of the 5 dimensions (data not shown).

## DISCUSSION

Our study investigated for the first time in the literature the role of the ASSR as a predictor of the response to antipsychotic medications, and it is one of the very few observing patients from the standpoint of an acute psychotic decompensation.

Results indicate that patients responding to treatment have reduced gamma band activity with high effect size (d=0.936), involving power (i.e. ERSP) but not phase synchrony (i.e. ITC). ERSP was also able to classify responders and non-responders with a 70% corrected accuracy, making it one of the stronger predictive biomarkers in psychiatry to date. Moreover, from a theoretical point of view, the relationship of ASSR parameters with the outcome of AP treatment corroborates the relevance of the ASSR-related processes for the pathophysiology of schizophrenia.

We observed that only responders to treatment had a reduced ERSP compared to controls, but not non-responders, and that no significant differences in ITC gamma emerged. These results at first glance contradict the literature ^14, 17^, which shows decreased ERSP and ITC in schizophrenia, with average effect size (g=-0.58 for ERSP and -0.46 for ITC), and only few studies finding no differences in patients vs controls ^22-26^, or an increased gamma power (at the level of the superior temporal gyrus ^27^). A more in-depth evaluation reveals instead a substantial coherence between our results and the literature: patients entering past studies were typically outpatients stabilized under an effective antipsychotic treatment, thus similar to responders to treatment in our cohort. Non-responders in the present study could represent a different phenotype, without significant gamma band impairment during the acute phase: we hypothesize that two distinct schizophrenia neurobiologies exist, one characterized by a significant E/I imbalance and an impairment in the PING microcircuits, the other relying on separate, so far unexplored, mechanisms.

Results, provocatively, questions the use of antipsychotic medications in patients not having a significant gamma band imbalance, which could be unable to benefit from an antipsychotic medication.

Only two previous study exist dealing with antipsychotic treatment and ASSR, though not designed to define predictors, but being a comparison of stabilized outpatients. First, Ogyu compared patients with or without treatment resistant schizophrenia (TRS, i.e.: the failure to respond to ≥ 2 different non-clozapine antipsychotic medications), showing that ERSP was lower in TRS compared to controls, but not in non-TRS compared to controls; the categorization of patients (i.e. TRS vs non-TRS) and the phase of the disease (i.e. not acute) were not overlapping to the ours, making results difficult to compare^28^. Second, Alegre compared acute drug naïve and well-stabilized patients under medication, concluding that antipsychotic medications can rescue gamma-band activity in patients with schizophrenia. In fact, drug-naive patients had reduced amplitude and inter-trial phase coherence of the response in the 30–50 Hz range, and reduced amplitude of the response in the 90–100 Hz range, when compared to controls. In the treated group, the response in the 30–50 Hz range was normalized to values similar to the control group^29^. The medication-induced recovery of a physiological level of gamma activity during the ASSR is a hypothesis not supported by the rest of the literature, considering that most of the patients entering previous studies were treated and well stabilized, yet showing a consistent decrease in ERSP and ITC. Moreover, in our study the effect size of the reduction in ERSP in responders to treatment during the acute phase is overlapping to the one observed in chronic, treated, outpatients, (Cohen’s d ∼ 0.5). Findings thus suggest that ERSP is a marker of the disease, remaining stable in different contexts, and not the direct expression of a pathophysiological process influenced by medications.

Previous investigations exist on EEG-based prediction of response to antipsychotics, mostly focused on resting state power spectra, thus being not directly comparable with our task-related results. Only two investigations produced evidence on resting-state gamma frequencies, indicating that both a reduced^30^ or an increased^31^ gamma power predicts a better response, with a third study addressing the issue and finding no significant results^32^.

The amount of clinical and preclinical evidence demonstrating the relevance of gamma activity for the pathophysiology of schizophrenia^7^, could favor ASSR as a potential biomarker in face of described alternatives without a strong biological rationale.

Most of the previous studies only analyzed a single or a limited number of medications, making results from one study to another difficult to compare. While our study cannot guide in the individualized choice of antipsychotic medications, we observed a consistency across specific medications categories, indicating that we individuated a general pattern of response to antipsychotics.

In contrast with the literature, we observed no differences between patients and controls for the ITC in gamma band; our interpretation is that ITC impairment is mainly a marker of the chronic-stabilized phase of schizophrenia. Accordingly, other studies not finding differences in ITC between patients and controls were the ones including patients with a higher severity of the disease, as indicated by the notion of an acute psychotic episode in the inpatients setting^29^, or of TRS and a PANSS score >90^28^. Interesting findings of our study, not being a focus of previous investigations, involved the theta band, including both ERSP and ITC. While not differentiating responders and non-responders to treatment, patients vs controls showed a reduction involving both the early and the late windows.

These results highlight the importance of theta band activity in schizophrenia, which was increased during the resting-state in chronic schizophrenia^33-35^, and related to reduced cognitive control/inhibition^36^, and to a more severe cognitive impairment^37, 38^, especially involving working memory and attention^39^. Thus, theta dysregulation in schizophrenia seems to consist of an increased baseline activity (as for past literature) and of a meager induced activity (as for our findings).

The functional relevance of theta band in schizophrenia and during the ASSR stimulation is further corroborated by our findings of an increased PAC in patients compared to controls, in both the early and the late phase. Our study is one of the few investigating PAC in schizophrenia, following investigations with negative results^18-20^ and one study showing a complex picture, with reduced PAC at the frontocentral, right middle temporal, and left temporoparietal electrodes but increased at the left parietal electrodes^21^. Our results suggest that instead of a loss of brain activity coherence, it is an excess of inflexibility and repetitiveness in the processes orchestrated by the theta-gamma coupling that could lead to aberrant salience and psychosis. Overall, some features overlapped with previous findings on stabilized outpatients and other differed, configuring a complex picture of trait markers of schizophrenia, stable across phases of the disease, and state markers of the acute psychosis or the chronic/stabilized phase of the disease. Our study also assessed the correlation of ASSR dynamics in ERSP, ITC and PAC with specific symptoms dimensions, finding no significant associations, in line with most previous studies. ITC was previously related to the severity of positive symptoms, however findings were contradictory, with a positive^40^ and a negative association^28^ concerning overall positive symptoms, and a negative association specifically with hallucinations^41^. Also, a single association was found between negative symptoms severity and the ERSP at 80 Hz^26^. Concerning PAC, the lack of correlations with symptoms dimensions aligns with past literature, with only one study indicating a negative correlation with working memory^18^.

It seems thus reasonable to conclude that ASSR dynamics mostly correlate with the pathophysiology process underlying schizophrenia as a whole entity, and only to a minor extent to each symptom dimension. We consider the lack of association particularly relevant, because we believe that our study on acute psychosis created the appropriate context to make such associations emerge, due to severity and heterogeneity of symptoms.

## Limitations

The sample size was limited, although if among largest to date for a study on the topic of response to antipsychotics with EEG and in the field of ASSR and schizophrenia. On one hand the sample size allows to establish a solid association between ASSR dynamics and response to medications; on another hand it is not sufficient to validate an established predictive biomarker.

Some of our methodological choices differed from previous investigations. First, we decided to split the ASSR into two segments, believing this is the more appropriate approach to appreciate the peculiar functional nature of the early, “ERP like” phase, and the late “ERSP proper” phase^15^. A common approach in literature is to consider one single window from stimulus onset to offset. Second, we chose to selectively assess the 40 Hz response, being the one more related to the resonance properties of the ASSR stimulus, while other studies used a broader signal^14, 17^. Third, factors associated with a stronger ASSR concerning power and phase are the younger age, the lower stimulus duration and the attentional load. This could explain a weaker effect in our study, where the average age was higher, the stimulus duration long and no active engagement of the patient was required, to permit acute patients to participate more easily^14^.

We had no EEG recording during the follow-up visit, which could have helped better clarifying the changing EEG dynamics corresponding to the process underlying response. However this choice is in line with the focus of the paper on prediction of response.

Another inherent limitation of the study is the lack of a long term assessment of a sustained response to medication; however the design of our study was aligned with the normal duration of clinical trials Patients were included within 72h of hospital admission, having already received initial doses of treatment. This approach significantly improved feasibility and we believe it had minimal potential for bias, as evidence suggests that 6–10 days are required for antipsychotic effects to become clinically significant^42-44^.

## CONCLUSIONS

The ASSR holds the potential to become predictive biomarker of response to antipsychotic medications, being to date the one with the highest effect size for discriminating responders and non-responders to treatment. Besides, our results suggest that responders and non-responders to treatment correspond to two different biotypes: only the former would be affected by a significant E/I imbalance, while in the latter a different pathophysiology would develop.

## Data Availability

All data produced in the present study are available upon reasonable request to the authors

