## Supplementary materials for "The 40 Hz auditory steady state response is associated with antipsychotic treatment outcome in acute patients with schizophrenia"

**Supplementary methods**
*Participants*
Adult inpatients (18–65 years) with schizophrenia were recruited within 72 hours of admission to an acute general psychiatric unit in Geneva, Switzerland. Diagnosis adhered to DSM-5 criteria, and inclusion required an acute psychotic episode, defined by the presence of delusions, hallucinations, and/or disorganization, alongside a Clinical Global Impression score of 4 or higher. Detailed inclusion and exclusion criteria are available in **Supplementary Table S1**.
Patients were twice evaluated clinically: upon EEG recording and again after six weeks. During this period, all patients received standard psychosocial treatment, including medical and nursing care, as well as social assistance when necessary. Clinical assessments were conducted by a senior psychiatrist through semi-structured interviews, clinical record reviews, and information gathered from caregivers and other relevant individuals. Psychopathology was assessed using the Positive and Negative Syndrome Scale^1^. Disease severity was evaluated by both the total PANSS score and its five symptom dimensions: positive (P1, P3, P5, G9), negative (N1, N2, N3, N4, N6, G7), excited (P4, P7, G8), disorganized (P2, N5, G11), and depressed (G2, G3, G6), following the framework by Wallwork et al.^2^.
All patients received antipsychotic medication from one of five categories: first-generation, -pines (serotonin and dopamine antagonists with prevalent antiserotoninergic activity), -dones (serotonin and dopamine antagonists with prevalent antidopaminergic activity), dopamine partial agonists (third generation antipsychotics, TGA), or amisulpride^3^. Adherence to treatment was confirmed by verifying the regular administration of long-acting injectable medication or through drug monitoring at the second evaluation, ensuring plasmatic concentrations were within the therapeutic interval. Response to antipsychotic treatment was defined as a 50% reduction in the total Positive and Negative Syndrome Scale (PANSS)^1^ score between the two evaluations.
A healthy control group was included, matched to patients by age, sex, handedness, and parental education level. Parental education was chosen over patient education to better account for the cognitive trajectory in schizophrenia^4^. Education levels were defined based on the Swiss education system legislation [https://www.edk.ch/en/education-system-ch]. The study was approved by the Ethics Committee of the Geneva Canton (ID 2023–0535). Informed consent was obtained from all participants, in accordance with national regulations on observational research in acute settings.

*ASSR procedure*

The task was administrated through a computer screen and in-ear earphones using Eprime software v3.0. While a white fixation cross was displayed on a black background, modulated and flat sounds were played. Modulated sound consisted of a 1000Hz pure tone sound modulated in amplitude from minus infinite to 0 dB with a 40Hz sinusoidal envelope, and flat sounds consisted of a 1000Hz pure tone. A total of 100 modulated and 10 flat sounds were played in pseudo random order (1 flat out of 11 sounds). Intertrial interval lengths were randomly picked between 0.5 and 2 sec.

*EEG acquisition and preprocessing*
EEG was recorded using an ANT Neuro EEGO MyLab system with 2 amplifiers and a 128-channel saline-soaked Waveguard EEG net. While the EEG net was installed and connected to the amplifiers, participants were seated in a calm room next to the experimenter. The hardware signal was referenced to Cz-corresponding electrode and recorded at a sampling rate of 500 Hz. All EEG analyses were performed in Python using MNE-Python v1.7. A channel for the reference electrode Cz was added offline. The EEG recorded during the task was first filtered with a band pass filter (FIR) between 1 and 80 Hz and a 50 Hz notch filter. Bad epochs and bad channels were visually marked as such prior to an ICA decomposition of the signal using the Picard method (Ablin et al., 2018). The ICA components corresponding to identifiable artifacts (i.e. eye blinks, saccades, ECG, facial muscle contractions) were discarded from the reconstructed signal. Bad channels were then interpolated based on neighbouring electrodes using the minimum-norm method, while bad epochs were discarded from further analysis. On average 8.46 ± 8.91 components were discarded from the reconstructed signal, and 7.42 ± 10.45 channels were interpolated The clean EEG was cut into epochs from -1 to 1.5 seconds related to the onset of the modulated sounds and 66.5±16 epochs were kept for each subject

*Time-frequency representation*
From these epochs, the average time-frequency representation (TFR) using Discrete Prolate Spheroidal Sequences (DPSS) tapers (MNE function tfr_multitaper, n_cycles = freqs / 2, use_fft = True, return_itc = True) together with the inter-trial coherence (ITC) were calculated initially from 4 to 80Hz with a 1-Hz resolution. For the surface analysis, the computation focused on a fronto-central cluster of 24 electrodes (Z4–Z9, L4–L9, R4–R9, LL5–LL8, RR5–RR8). An optimal 0.25-s baseline was individually identified within −0.9–0 s as the window showing maximal contrast with the late-ASSR period (0.3–1.2 s), and this baseline was applied using log-ratio correction. From the baseline-corrected TFRs, mean event-related spectral perturbation (ERSP) and inter-trial phase coherence (ITPC) were extracted in two frequency bins and different time windows defined a priori: theta (4-8 Hz; 0–0.5 s window) and gamma (40 Hz; early 0–0.3 s, late 0.3–1.2 s). For each subject and electrode, ERSP and ITPC metrics were averaged across time, categorising ASSR by time windows and frequency bins.
For statistical evaluation, subject-level ERSP and ITPC metrics derived from the theta (4-8 Hz) and gamma (40 Hz) bins (two time windows) were analysed in groups according to clinical metadata for diagnostic and treatment-response i.e., controls, patients, responders, non-responders, then organized into four planned group comparisons: patients vs. controls, responders vs. non-responders, responders vs. controls, and non-responders vs. controls. Missing values were replaced with zeros.

*Phase amplitude coupling*
Phase–amplitude coupling (PAC) was also computed for baseline, early, and late temporal windows for each electrode using a modulation index (MI) approach based on the Kullback–Leibler divergence of the amplitude–phase distribution. For each ASSR epoch, low-frequency phase (4–8 Hz) and high-frequency amplitude (39–41 Hz) components were extracted via band-pass filtering of the EEG signal and analytic signal estimation with the Hilbert transform. Instantaneous phase and amplitude envelopes were binned into 18 uniform phase intervals (−π to π rad), and the mean amplitude within each phase bin was used to derive the phase–amplitude distribution. The modulation index (MI) was then calculated as the normalized difference between the observed and uniform entropy of this distribution. MI values were averaged across epochs to obtain a single PAC estimate per electrode and analysis window. Two temporal windows were analyzed: early (0–0.2 s) and late (0.2–1.2 s).

*Statistics*
We compared the three study groups (Responders, Non-Responders, and Controls) across key demographic and clinical variables, including age, sex, handedness, education level (subject and parents), age of onset, illness duration, duration of untreated psychosis (DUP), and number of hospitalizations. Clinical symptoms were assessed using PANSS scores (Total, Positive, Negative, Disorganized, Excited, Depressed). Additionally, Responders and Non-Responders were specifically compared regarding their antipsychotic medication regimens, including medication status before admission, antipsychotic category, specific agents (first and second antipsychotic), presence of polypharmacy, and chlorpromazine equivalent doses. Statistical comparisons employed Chi-Square or Fisher’s Exact tests for categorical variables (reporting Cramer’s V) and Mann-Whitney U tests for continuous variables (reporting Cohen’s d).
To assess potential confounding effects, we analyzed associations between clinical variables (e.g., age, CPZ equivalents) and electrophysiological metrics. For ERSP, ITPC, and PAC alike, data were averaged across the 25 ROI electrodes to obtain a single representative value per subject for each time window (Early, Late) and frequency band (where applicable). Statistical dependence was evaluated using Spearman’s rank correlation for continuous variables and Kruskal-Wallis tests for categorical factors, with $p$-values corrected for multiple comparisons using the Benjamini-Hochberg FDR procedure.
For each band (theta, gamma), time window (early, late) and electrophysiological metric (ERSP, ITC, PAC), group differences (responders vs non-responders, patients vs controls, responders vs controls, non responders vs controls) were then assessed using non-parametric cluster-based permutation tests implemented in MNE-Python, which identifies clusters of electrodes showing significant differences between groups, controlling for multiple comparisons via the cluster-wise permutation approach. Each test used 10,000 random permutations, a one-tailed design for ERSP and ITC, and two-tailed for PAC, and a spatial adjacency matrix. Clusters with corrected p < 0.05 were considered significant. For significant cluster (p-value < 0.05), a Cohen's d effect size was calculated (absolute d >= 0.8 "Large"; d >= 0.5 "Medium"; d >= 0.2 "Small"; d < 0.02 "Very Small"), in order to investigate and identify the main effects across the different comparisons.
A correlational analysis between ASSR features and PANSS subscales for positive, negative, disorganized, excited and depressed symptoms was performed, using a cluster based permutation test, using the same parameters described for group comparisons. The statistical test was defined as the *t*-value derived from the Pearson correlation coefficient ($r$) between the EEG data and the outcome variable.

*Prediction of Treatment Response and Internal Validation*
To evaluate the translational utility of the identified electrophysiological deficits, we constructed a binary logistic regression model to predict clinical treatment response (Responder vs. Non-Responder) based on the ASSR features. To avoid feature selection bias and data leakage, the predictor variable was derived strictly from the results of the spatial cluster-based permutation test. We calculated the mean ERSP value across the specific subset of electrodes forming the statistically significant spatial cluster that differentiated Responders from Non-Responders. This cluster-averaged value served as the continuous predictor variable, while the clinical response status (0/1) served as the binary outcome. The predictor was standardized (z-scored) prior to model fitting to ensure convergence. Model performance was evaluated using Receiver Operating Characteristic (ROC) curve analysis. The Area Under the Curve (AUC) was calculated to quantify discriminative ability. To convert the model’s continuous probability outputs into binary class predictions, we identified the optimal classification threshold using Youden’s Index ($J=Sensitivity+Specificity-1$), which maximizes the balance between sensitivity and specificity. Given the sample size and the risk of overfitting (optimism) associated with training and testing on the same dataset, we applied Harrell’s bootstrap optimism correction method to validate the results. We performed 1,000 bootstrap iterations. In each iteration, a bootstrap sample was drawn with replacement from the original dataset, stratified by response status, the logistic regression model was fitted to this bootstrap sample, the model’s performance (AUC, Accuracy, Sensitivity, Specificity) was evaluated on the bootstrap sample itself ("bootstrap performance") and subsequently on the original dataset ("test performance"); finally, the difference between the bootstrap performance and test performance was recorded as the "optimism." The average optimism across all 1,000 iterations was subtracted from the apparent performance metrics of the original model to yield bias-corrected estimates.

*References*
**1.**	Kay SR, Fiszbein A, Opler LA. The positive and negative syndrome scale (PANSS) for schizophrenia. *Schizophr Bull.* 1987;13(2):261-276.

**2.**	Wallwork RS, Fortgang R, Hashimoto R, Weinberger DR, Dickinson D. Searching for a consensus five-factor model of the Positive and Negative Syndrome Scale for schizophrenia. *Schizophr Res.* May 2012;137(1-3):246-250.

**3.**	Stahl SM. Beyond the dopamine hypothesis of schizophrenia to three neural networks of psychosis: dopamine, serotonin, and glutamate. *CNS Spectr.* Jun 2018;23(3):187-191.

**4.**	McCutcheon RA, Reis Marques T, Howes OD. Schizophrenia-An Overview. *JAMA Psychiatry.* Feb 1 2020;77(2):201-210.

**Supplementary table S1:** inclusion and exclusion criteria for participation in the study

| *Inclusion criteria (for patients with schizophrenia)*   - A diagnosis of schizophrenia according to the diagnostic and statistical manual of mental disorders (DSM5) - A clinical global impression (CGI) score ≥ 4 pts (“moderately ill” or worse) - Indication to an antipsychotic treatment - Informed consent as documented by signature | *Exclusion criteria (for patients with schizophrenia)*   - Age below 18 and beyond 65 years - Concomitant substance use disorder, according to DSM5 criteria - Concomitant medication with antidepressant and/or mood stabilizers - Other clinically significant concomitant disease states (e.g., renal failure, hepatic dysfunction, cardiovascular disease, major neurological disorders) - Known or suspected non-compliance to treatment in the inpatient setting   *Exclusion criteria (for healthy control)*   - History of mental illness - Concomitant substance use disorder, according to DSM5 criteria - Other clinically significant concomitant disease states (e.g., renal failure, hepatic dysfunction, cardiovascular disease, major neurological disorders) |
| --- | --- |

**Supplementary table S2**: potential associations of confounders with ERSP, ITC, PAC

| Metric | Confounder | Test Type | Coef (r/η) | Raw P | FDR P |
| --- | --- | --- | --- | --- | --- |
| ERSP early Gamma | Age | Rho (Spearman) | -0.116 | 0.2225 | 0.8900 |
|  | Sex | Eta (Kruskal) | 0.000 | 0.7433 | 0.9288 |
|  | Handedness | Eta (Kruskal) | 0.000 | 0.5460 | 0.9288 |
|  | Education | Rho (Spearman) | 0.020 | 0.8312 | 0.9288 |
|  | Education mother | Rho (Spearman) | -0.051 | 0.6410 | 0.9288 |
|  | Education father | Rho (Spearman) | -0.021 | 0.8514 | 0.9288 |
|  | Age of onset | Rho (Spearman) | -0.119 | 0.3248 | 0.9288 |
|  | Duration of illness | Rho (Spearman) | -0.006 | 0.9621 | 0.9621 |
|  | Duration of untreated psychosis | Rho (Spearman) | 0.249 | 0.0617 | 0.4291 |
|  | Number of hospitalizations | Rho (Spearman) | -0.047 | 0.6899 | 0.9288 |
|  | Risperidone equivalents | Rho (Spearman) | -0.098 | 0.4041 | 0.9288 |
|  | Antipsychotic category | Eta (Kruskal) | 0.257 | 0.0715 | 0.4291 |
| ERSP late Gamma | Age | Rho (Spearman) | -0.151 | 0.1103 | 0.8935 |
|  | Sex | Eta (Kruskal) | 0.000 | 0.7537 | 0.8935 |
|  | Handedness | Eta (Kruskal) | 0.000 | 0.4939 | 0.8935 |
|  | Education | Rho (Spearman) | -0.038 | 0.6947 | 0.8935 |
|  | Education mother | Rho (Spearman) | -0.050 | 0.6479 | 0.8935 |
|  | Education father | Rho (Spearman) | -0.063 | 0.5669 | 0.8935 |
|  | Age of onset | Rho (Spearman) | -0.039 | 0.7506 | 0.8935 |
|  | Duration of illness | Rho (Spearman) | -0.023 | 0.8554 | 0.8935 |
|  | Duration of untreated psychosis | Rho (Spearman) | 0.066 | 0.6241 | 0.8935 |
|  | Number of hospitalizations | Rho (Spearman) | -0.073 | 0.5414 | 0.8935 |
|  | Risperidone equivalents | Rho (Spearman) | 0.016 | 0.8935 | 0.8935 |
|  | Antipsychotic category | Eta (Kruskal) | 0.133 | 0.2644 | 0.8935 |
| ERSP early Theta | Age | Rho (Spearman) | -0.094 | 0.3245 | 0.6142 |
|  | Sex | Eta (Kruskal) | 0.000 | 0.4654 | 0.6161 |
|  | Handedness | Eta (Kruskal) | 0.092 | 0.2479 | 0.5972 |
|  | Education | Rho (Spearman) | 0.250 | 0.0101 | 0.0701 |
|  | Education mother | Rho (Spearman) | 0.212 | 0.0499 | 0.2997 |
|  | Education father | Rho (Spearman) | 0.126 | 0.2488 | 0.5972 |
|  | Age of onset | Rho (Spearman) | -0.075 | 0.5393 | 0.6161 |
|  | Duration of illness | Rho (Spearman) | 0.034 | 0.7865 | 0.7865 |
|  | Duration of untreated psychosis | Rho (Spearman) | 0.168 | 0.2128 | 0.5972 |
|  | Number of hospitalizations | Rho (Spearman) | -0.068 | 0.5648 | 0.6161 |
|  | Risperidone equivalents | Rho (Spearman) | -0.108 | 0.3583 | 0.6142 |
|  | Antipsychotic category | Eta (Kruskal) | 0.000 | 0.4836 | 0.6161 |
| ERSP late Theta | Age | Rho (Spearman) | -0.142 | 0.1346 | 0.8075 |
|  | Sex | Eta (Kruskal) | 0.000 | 0.9674 | 0.9674 |
|  | Handedness | Eta (Kruskal) | 0.054 | 0.3218 | 0.8202 |
|  | Education | Rho (Spearman) | 0.173 | 0.0692 | 0.8075 |
|  | Education mother | Rho (Spearman) | 0.067 | 0.5412 | 0.8202 |
|  | Education father | Rho (Spearman) | 0.074 | 0.5022 | 0.8202 |
|  | Age of onset | Rho (Spearman) | -0.124 | 0.3057 | 0.8202 |
|  | Duration of illness | Rho (Spearman) | 0.075 | 0.5468 | 0.8202 |
|  | Number of hospitalizations | Rho (Spearman) | 0.022 | 0.8554 | 0.9674 |
|  | Duration of untreated psychosis | Rho (Spearman) | 0.120 | 0.3746 | 0.8202 |
|  | Antipsychotic category | Eta (Kruskal) | 0.000 | 0.8150 | 0.9674 |
|  | Risperidone equivalents | Rho (Spearman) | -0.015 | 0.8961 | 0.9674 |
| ITC early Gamma | Age | Rho (Spearman) | -0.153 | 0.1060 | 0.3658 |
|  | Sex | Eta (Kruskal) | 0.000 | 0.4489 | 0.5865 |
|  | Handedness | Eta (Kruskal) | 0.000 | 0.7409 | 0.7409 |
|  | Education | Rho (Spearman) | -0.185 | 0.0213 | 0.1421 |
|  | Education mother | Rho (Spearman) | -0.168 | 0.1219 | 0.3658 |
|  | Education father | Rho (Spearman) | -0.068 | 0.5376 | 0.5865 |
|  | Age of onset | Rho (Spearman) | -0.161 | 0.1836 | 0.4406 |
|  | Duration of illness | Rho (Spearman) | -0.084 | 0.4983 | 0.5865 |
|  | Duration of untreated psychosis | Rho (Spearman) | -0.089 | 0.5081 | 0.5865 |
|  | Number of hospitalizations | Rho (Spearman) | -0.086 | 0.4694 | 0.5865 |
|  | Antipsychotic category | Eta (Kruskal) | 0.148 | 0.2370 | 0.4741 |
|  | Risperidone equivalents | Rho (Spearman) | 0.263 | 0.0224 | 0.1343 |
| ITC late Gamma | Age | Rho (Spearman) | -0.151 | 0.1105 | 0.4419 |
|  | Sex | Eta (Kruskal) | 0.000 | 0.3642 | 0.6759 |
|  | Handedness | Eta (Kruskal) | 0.000 | 0.6948 | 0.8337 |
|  | Education | Rho (Spearman) | -0.210 | 0.0500 | 0.0912 |
|  | Education mother | Rho (Spearman) | -0.089 | 0.4168 | 0.6759 |
|  | Education father | Rho (Spearman) | -0.048 | 0.6612 | 0.8337 |
|  | Age of onset | Rho (Spearman) | -0.205 | 0.0884 | 0.4419 |
|  | Duration of illness | Rho (Spearman) | -0.026 | 0.8342 | 0.8720 |
|  | Duration of untreated psychosis | Rho (Spearman) | 0.022 | 0.8720 | 0.8720 |
|  | Number of hospitalizations | Rho (Spearman) | -0.123 | 0.2989 | 0.6759 |
|  | Antipsychotic category | Eta (Kruskal) | 0.000 | 0.4506 | 0.6759 |
|  | Risperidone equivalents | Rho (Spearman) | 0.158 | 0.1750 | 0.5250 |
| ITC early Theta | Age | Rho (Spearman) | 0.062 | 0.5135 | 0.7988 |
|  | Sex | Eta (Kruskal) | 0.000 | 0.6137 | 0.7988 |
|  | Handedness | Eta (Kruskal) | 0.000 | 0.5542 | 0.7988 |
|  | Education | Rho (Spearman) | 0.215 | 0.0231 | 0.2774 |
|  | Education mother | Rho (Spearman) | 0.170 | 0.1166 | 0.6994 |
|  | Education father | Rho (Spearman) | 0.048 | 0.6657 | 0.7988 |
|  | Age of onset | Rho (Spearman) | 0.096 | 0.4272 | 0.7988 |
|  | Duration of illness | Rho (Spearman) | 0.001 | 0.9950 | 0.9950 |
|  | Duration of untreated psychosis | Rho (Spearman) | 0.083 | 0.5374 | 0.7988 |
|  | Number of hospitalizations | Rho (Spearman) | -0.016 | 0.8919 | 0.9730 |
|  | Antipsychotic category | Eta (Kruskal) | 0.157 | 0.2209 | 0.7988 |
|  | Risperidone equivalents | Rho (Spearman) | 0.061 | 0.6015 | 0.7988 |
| ITC late Theta | Age | Rho (Spearman) | -0.040 | 0.6749 | 0.7362 |
|  | Sex | Eta (Kruskal) | 0.209 | 0.0157 | 0.1889 |
|  | Handedness | Eta (Kruskal) | 0.000 | 0.6615 | 0.7362 |
|  | Education | Rho (Spearman) | 0.082 | 0.3901 | 0.7265 |
|  | Education mother | Rho (Spearman) | 0.138 | 0.2067 | 0.7265 |
|  | Education father | Rho (Spearman) | 0.135 | 0.2166 | 0.7265 |
|  | Age of onset | Rho (Spearman) | -0.060 | 0.6201 | 0.7362 |
|  | Duration of illness | Rho (Spearman) | 0.117 | 0.3471 | 0.7265 |
|  | Duration of untreated psychosis | Rho (Spearman) | -0.044 | 0.7465 | 0.7465 |
|  | Number of hospitalizations | Rho (Spearman) | 0.077 | 0.5172 | 0.7362 |
|  | Antipsychotic category | Eta (Kruskal) | 0.139 | 0.2523 | 0.7265 |
|  | Risperidone equivalents | Rho (Spearman) | 0.094 | 0.4238 | 0.7265 |
| PAC Early | Antipsychotic category | Eta (Kruskal) | 0.294 | 0.0425 | 0.4770 |
|  | Risperidone equivalents | Rho (Spearman) | 0.158 | 0.1823 | 0.7294 |
|  | Age | Rho (Spearman) | 0.068 | 0.4866 | 0.8011 |
|  | Education | Rho (Spearman) | -0.172 | 0.0795 | 0.4770 |
|  | Sex | Eta (Kruskal) | 0.000 | 0.6008 | 0.8011 |
|  | Education mother | Rho (Spearman) | -0.074 | 0.5188 | 0.8011 |
|  | Education father | Rho (Spearman) | 0.079 | 0.4878 | 0.8011 |
|  | Age of onset | Rho (Spearman) | -0.065 | 0.5982 | 0.8011 |
|  | Duration of untreated psychosis | Rho (Spearman) | -0.084 | 0.5417 | 0.8011 |
|  | Handedness | Eta (Kruskal) | 0.000 | 0.8144 | 0.8173 |
|  | Duration of illness | Rho (Spearman) | 0.029 | 0.8173 | 0.8173 |
|  | Number of hospitalizations | Rho (Spearman) | 0.039 | 0.7456 | 0.8173 |
| PAC Late | Antipsychotic category | Eta (Kruskal) | 0.200 | 0.0212 | 0.0621 |
|  | Risperidone equivalents | Rho (Spearman) | 0.290 | 0.0129 | 0.0531 |
|  | Duration of illness | Rho (Spearman) | 0.293 | 0.0177 | 0.0531 |
|  | Number of hospitalizations | Rho (Spearman) | 0.285 | 0.0159 | 0.0531 |
|  | Education | Rho (Spearman) | -0.221 | 0.0238 | 0.0571 |
|  | Sex | Eta (Kruskal) | 0.170 | 0.0445 | 0.0889 |
|  | Handedness | Eta (Kruskal) | 0.147 | 0.1442 | 0.2163 |
|  | Age of onset | Rho (Spearman) | -0.182 | 0.1380 | 0.2163 |
|  | Education mother | Rho (Spearman) | -0.155 | 0.1735 | 0.2313 |
|  | Age | Rho (Spearman) | 0.111 | 0.2554 | 0.3065 |
|  | Duration of untreated psychosis | Rho (Spearman) | 0.023 | 0.8668 | 0.9456 |
|  | Education father | Rho (Spearman) | -0.003 | 0.9796 | 0.9796 |

**Supplementary table S3**: ERSP comparisons between responders, non-responders and controls

|  | Electrodes (Cluster) | p-value | ES (d) |  | |
| --- | --- | --- | --- | --- | --- |
| Responders vs non-responders | | | |  |  |
|  |  |  |  | *Responders* | *Non-responders* |
| ERSP early 4-8 Hz | R7, R8, RR6 | 0.0472 | -0.677 | 0.051±0.077 | 0.097±0.055 |
| ERSP late 4-8 Hz | Z9, R8, R9, RR8 | 0.0287 | -0.676 | 0.058±0.068 | 0.102±0.06 |
| ERSP early 40 Hz | Z4, Z5, Z6, Z8, Z9, L6, L7, L9, R5, R6, R7, R8, LL8, RR6 | 0.0013 | -0.936 | 0.069±0.035 | 0.1±0.032 |
| ERSP late 40 Hz | Z4, Z5, Z6, Z8, Z9, L6, L7, L9, R5, R6, R7, R8, RR5, RR6 | 0.0022 | -0.932 | 0.071±0.042 | 0.106±0.033 |
| Responders vs controls | | | |  | |
|  |  |  |  | *Responders* | *Controls* |
| ERSP early 4-8 Hz | R8, RR7, RR8 | 0.0337 | -0.649 | 0.07±0.079 | 0.124±0.09 |
| ERSP late 4-8 Hz | n.s. (Global Avg) | 0.3266 | -0.087 | 0.073±0.048 | 0.078±0.077 |
| ERSP early 40 Hz | n.s. (Global Avg) | 0.1672 | -0.221 | 0.07±0.036 | 0.078±0.037 |
| ERSP late 40 Hz | Z8, Z9, L7, L9, R8, LL6, LL8 | 0.0211 | -0.558 | 0.064±0.055 | 0.093±0.045 |
| Non-responders vs controls | | | |  | |
|  |  |  |  | *Non responders* | *Controls* |
| ERSP early 4-8 Hz | n.s. (Global Avg) | 0.0514 | -0.256 | 0.099±0.056 | 0.117±0.082 |
| ERSP late 4-8 Hz | n.s. (Global Avg) | 1.0000 | 0.207 | 0.092±0.052 | 0.078±0.077 |
| ERSP early 40 Hz | n.s. (Global Avg) | 1.0000 | 0.461 | 0.093±0.031 | 0.078±0.037 |
| ERSP late 40 Hz | n.s. (Global Avg) | 1.0000 | 0.243 | 0.098±0.032 | 0.089±0.044 |
| Patients vs controls | | | |  | |
|  |  |  |  | *Patients* | *Controls* |
| ERSP early 4-8 Hz | Z4, R4, R5 | 0.0390 | -0.585 | 0.092±0.065 | 0.134±0.084 |
| ERSP late 4-8 Hz | n.s. (Global Avg) | 1.0000 | -0.024 | 0.077±0.055 | 0.078±0.077 |
| ERSP early 40 Hz | n.s. (Global Avg) | 0.2028 | 0.03 | 0.079±0.038 | 0.078±0.037 |
| ERSP late 40 Hz | n.s. (Global Avg) | 0.3305 | -0.132 | 0.083±0.042 | 0.089±0.044 |

**Supplementary table S4**: ITC comparisons between responders, non-responders and controls

|  | Electrodes (Cluster) | p-value | Cohen’s d |  | |
| --- | --- | --- | --- | --- | --- |
| Responders vs non-responders | | | |  |  |
|  |  |  |  | *Responders* | *Non-responders* |
| ITC early 4-8 Hz | n.s. (Global Avg) | 1.0000 | 0.028 | 0.184±0.056 | 0.182±0.063 |
| ITC late 4-8 Hz | n.s. (Global Avg) | 0.0885 | -0.138 | 0.125±0.024 | 0.128±0.019 |
| ITC early 40 Hz | n.s. (Global Avg) | 1.0000 | -0.087 | 0.161±0.043 | 0.165±0.057 |
| ITC late 40 Hz | n.s. (Global Avg) | 1.0000 | -0.223 | 0.163±0.044 | 0.173±0.054 |
| Responders vs controls | | | |  | |
|  |  |  |  | *Responders* | *Controls* |
| ITC early 4-8 Hz | Z4, Z5, Z6, Z8, Z9, L4, L5, L6, L8, L9, R4, R5, R6, R7, R8, R9, LL5, LL6, RR5, RR6, RR7, RR8 | <0.001 | -1.078 | 0.185±0.06 | 0.253±0.066 |
| ITC late 4-8 Hz | Z4, Z5, L5, L6, R4, R5, R6, LL5, RR5 | 0.0236 | -0.476 | 0.123±0.03 | 0.136±0.023 |
| ITC early 40 Hz | n.s. (Global Avg) | 1.0000 | 0.503 | 0.161±0.043 | 0.142±0.032 |
| ITC late 40 Hz | n.s. (Global Avg) | 1.0000 | 0.15 | 0.163±0.044 | 0.155±0.049 |
| Non-responders vs controls | | | |  | |
|  |  |  |  | *Non-responders* | *Controls* |
| ITC early 4-8 Hz | Z4, Z5, Z6, Z8, Z9, L4, L5, L6, L8, L9, R4, R5, R6, R7, R8, R9, LL5, LL6, LL8, RR5, RR6, RR7, RR8 | <0.001 | -1.04 | 0.184±0.064 | 0.251±0.065 |
| ITC late 4-8 Hz | n.s. (Global Avg) | 0.0667 | -0.262 | 0.128±0.019 | 0.133±0.017 |
| ITC early 40 Hz | n.s. (Global Avg) | 1.0000 | 0.517 | 0.165±0.057 | 0.142±0.032 |
| ITC late 40 Hz | n.s. (Global Avg) | 1.0000 | 0.348 | 0.173±0.054 | 0.155±0.049 |
| Patients vs controls | | | |  | |
|  |  |  |  | *Patients* | *Controls* |
| ITC early 4-8 Hz | Z4, Z5, Z6, Z8, Z9, L4, L5, L6, L8, L9, R4, R5, R6, R7, R8, R9, LL5, LL6, LL8, RR5, RR6, RR7, RR8 | <0.001 | -1.015 | 0.188±0.061 | 0.251±0.065 |
| ITC late 4-8 Hz | n.s. (Global Avg) | 0.0572 | -0.237 | 0.128±0.022 | 0.133±0.017 |
| ITC early 40 Hz | n.s. (Global Avg) | 1.0000 | 0.488 | 0.163±0.048 | 0.142±0.032 |
| ITC late 40 Hz | n.s. (Global Avg) | 1.0000 | 0.258 | 0.168±0.047 | 0.155±0.049 |

**Supplementary table S5**: PAC comparisons between responders, non-responders and controls

|  | Electrodes | P-Value | Cohen's d |  | |
| --- | --- | --- | --- | --- | --- |
| Responders vs non-responders | | | |  |  |
|  |  |  |  | *Responders* | *Non-responders* |
| PAC Early | Z4, L4, L5 | 0.02150 | 0.65 | 0.0006 ± 0.0005 | 0.0003 ± 0.0002 |
| PAC Late | Global Avg (Ns) | Ns | 0.21 | 0.0001 ± 0.0001 | 0.0001 ± 0.0000 |
| Responders vs controls | | | |  | |
|  |  |  |  | *Responders* | *Controls* |
| PAC Early | Z4, Z5, Z6, Z8, L4, L5, L6, L7, R4, LL6, RR5 | 0.00030 | 0.82 | 0.0005 ± 0.0004 | 0.0003 ± 0.0001 |
| PAC Late | Z8, Z9, R8, R9, RR8 | 0.00780 | 0.68 | 0.0002 ± 0.0001 | 0.0001 ± 0.0000 |
| Non-responders vs controls | | | |  | |
|  |  |  |  | *Non-responders* | *Controls* |
| PAC Early | Global Avg (Ns) | Ns | 0.60 | 0.0004 ± 0.0002 | 0.0003 ± 0.0001 |
| PAC Late | Z8, L7, L8, R7 | 0.01350 | 0.88 | 0.0001 ± 0.0001 | 0.0001 ± 0.0000 |
| Patients vs controls | | | |  | |
|  |  |  |  | *Patients* | *Controls* |
| PAC Early | Z4, Z5, Z6, Z8, L4, L5, L6, L7, R4, R5, R6, R9, LL6, RR5, RR6, RR7, RR8 | 0.00020 | 0.57 | 0.0005 ± 0.0004 | 0.0003 ± 0.0001 |
| PAC Late | Z4, Z6, Z8, Z9, L5, L6, L7, L8, R4, R6, R7, R8, R9, RR6, RR8 | 0.00060 | 0.74 | 0.0001 ± 0.0001 | 0.0001 ± 0.0000 |
